# The mental health impact of the covid-19 pandemic on healthcare workers, and interventions to help them: a rapid systematic review

**DOI:** 10.1101/2020.07.03.20145607

**Authors:** Ashley Elizabeth Muller, Elisabet Vivianne Hafstad, Jan Peter William Himmels, Geir Smedslund, Signe Flottorp, Synne Øien Stensland, Stijn Stroobants, Stijn van de Velde, Gunn Elisabeth Vist

## Abstract

**Background:** The covid-19 pandemic has heavily burdened, and in some cases overwhelmed, healthcare systems throughout the world. Healthcare workers are not only at heightened risk of infection, but also of adverse mental health outcomes. Identification of organizational, collegial and individual risk and resilience factors impacting the mental health of healthcare workers are needed to inform preparedness planning and sustainable response.

**Methods:** We performed a rapid systematic review to identify, assess and summarize available research on the mental health impact of the covid-19 pandemic on healthcare workers. On 11 May 2020, we utilized the Norwegian Institute of Public Health’s *Live map of covid-19 evidence*, the visualization of a database of 20,738 screened studies, to identify studies for inclusion. We included studies reporting on any type of mental health outcome in any type of healthcare workers during the pandemic. We described interventions reported by the studies, and narratively summarized mental health-related outcomes, as study heterogeneity precluded meta-analysis. We assessed study quality using design-specific instruments.

**Results:** We included 59 studies, reporting on a total of 54,707 healthcare workers. The prevalence of general psychological distress across the studies ranged from 7-97% (median 37%), anxiety 9-90% (median 24%), depression 5-51% (median 21%), and sleeping problems 34-65% (median 37%). Seven studies reported on implementing mental health interventions, and most focused on individual symptom reduction, but none reported on effects of the interventions. In most studies, healthcare workers reported low interest in and use of professional help, and greater reliance on social support and contact with family and friends. Exposure to covid-19 was the most commonly reported correlate of mental health problems, followed by female gender, and worry about infection or about infecting others. Social support correlated with less mental health problems.

**Discussion:** Healthcare workers in a variety of fields, positions, and exposure risks are reporting anxiety, depression, sleep problems, and distress during the covid-19 pandemic, but most studies do not report comparative data on mental health symptoms. before the pandemic. There seems to be a mismatch between risk factors for adverse mental health outcomes among healthcare workers in the current pandemic and their needs and preferences, and the individual psychopathology focus of current interventions. Efforts to help healthcare workers sustain healthy relationships to colleagues, family and friends over time may be paramount to safeguard what is already an important source of support during the prolonged crisis. Expanding interventions’ focus to incorporate organizational, collegial and family factors to support healthcare workers responding to the pandemic could improve acceptability and efficacy of interventions.

**Other:** The protocol for this review is available online. No funding was received.

Summary box

What is already known on this topic

- During viral outbreaks such as covid-19, healthcare providers are at increased risk of infection and negative physical and mental health outcomes
- Covid-19 is a particular challenge to healthcare systems and workers

What this study adds

- Healthcare workers’ mental health problems correlate with organizational factors such as workload and exposure to covid-19 patients
- Healthcare workers are more interested in occupational protection, rest, and social support than in professional psychological help
- Interventions focus more on addressing individual psychopathology, which points towards a mismatch between what workers want and need, and the services available to them

## Introduction

The covid-19 pandemic has heavily burdened, and in many cases overwhelmed, healthcare systems ^1, 2^ including healthcare workers. The WHO emphasized the extremely high burden on healthcare workers, and called for action to address the immediate needs and measures needed to save lives and prevent a serious impact on physical and mental health of healthcare workers ^3^.

Previous viral outbreaks have shown that frontline and non-frontline healthcare workers are at increased risk of infection and other adverse physical health outcomes ^4^. Furthermore, mental health problems putatively associated with healthcare workers’ occupational activities were reported during and up until years after epidemics, including symptoms of post-traumatic stress, burnout, depression and anxiety ^5-7^. Likewise, reports of the mental toll on Healthcare workers have persistently appeared during the current global health crisis ^8-10^.

Several reviews have already been conducted on healthcare workers’ mental health in the covid-19 pandemic, with search dates up to May 2020. Pappa et al. ^11^ identified thirteen studies in a search on 17 April 2020 and pooled prevalence rates; they reported that more than one of every five healthcare workers suffered from anxiety and/or depression; nearly two in five reported insomnia. Vindegaard & Benros’ ^12^ review, searching on 10 May 2020, identified twenty studies of healthcare workers in a subgroup analysis, and their narrative summary concluded that healthcare workers generally report more anxiety, depression, and sleep problems compared with the general population.

In the face of a prolonged crisis such as the pandemic, sustainability of the healthcare response fully relies on its ability to safeguard the health of responders: the healthcare workers^13, 14^. Yet, the recent findings of psychological distress among healthcare workers might indicate that the healthcare system is currently unable to effectively help the helpers. Understanding the risks and mental health impact(s) that healthcare workers experience, and identifying possible interventions to address adverse effects, is invaluable. Our main aim was to perform a rapid systematic review to identify, assess and summarize available research on the mental health impact of the covid-19 pandemic on healthcare workers and on healthcare workers’ understandings of their own mental health during the pandemic. Our second aim was to describe the interventions assessed in the literature to prevent or reduce negative mental health impacts on healthcare workers who are at work during the covid-19 pandemic.

## Method

We conducted a rapid systematic review according to the methods specified in our protocol, published on our institution’s website ^15^.

### Inclusion criteria

We included any type of study about any type of healthcare worker during the covid-19 pandemic, with outcomes relating to their mental health. We extracted information about interventions aimed at preventing or reducing negative mental health impacts on healthcare workers. We had no restrictions related to study design, methodological quality, or language.

### Literature search and article selection

We identified relevant studies by searching the Norwegian Institute of Public Health’s (NIPH’s) *Live map of covid-19 evidence* (https://www.fhi.no/en/qk/systematic-reviews-hta/map/) and database on 11 May 2020, as described in our protocol^15^. The live map and database contained 20,738 references screened for covid-19 relevance containing primary, secondary, or modelled data. Two researchers independently categorized these references according to topic (seven main topics, 52 subordinate topics), population (41 available groups), study design, and publication type. We identified references categorized to the population “*Healthcare worker*s”, and to the topic “*Experiences and perceptions, consequences; social, political, economic aspects*”. In addition, we identified references by searching (title/abstract) in the live map’s database, using the keywords: emo*, psych*, stress*, anx*, depr*, mental*, sleep, worry, somatoform, and somatic symptom disorder. We screened all identified references specifically for the inclusion criteria for this systematic review.

The protocol of the *Live map of covid-19 evidence* describes the methodology of the map and database ^16^. The methodology, including the search, has developed dynamically since March 2020. We performed our first search for the map 12.03.2020 and we have identified references published since 01.12.2019 by searching:

- PubMed (National Library of Medicine), from 01.12.2019 - 03.05.2020
- Embase (Ovid), between 01.12.2019 - 27.03.2020
- Centers for Disease Control and Prevention (CDC), 01.12.2019 - 11.05.2020

The last included search for this review was conducted on 11 May 2020. The search strategy is presented in Appendix 1.

### Data extraction and methodological quality assessment

We developed a data extraction form to collect data on country and setting, participants, exposure to covid-19, intervention if relevant, and outcomes related to mental health. We extracted data on prevalence of mental health problems as well as correlates (i.e. risk/resilience factors); strategies implemented or accessed by healthcare worker to address their own mental health; perceived need and preferences related to interventions aimed at preventing or reducing negative mental health consequences; and experience and understandings of mental health and related interventions. One researcher (AEM) extracted data and another checked her extraction. Two researchers independently assessed the methodological quality of systematic reviews using the AMSTAR tool ^17^ and of qualitative studies using the CASP checklist ^18^. One researcher (AEM, SF) assessed the quality of cross-sectional studies using either the JBI Prevalence or the JBI Cross-sectional Analytical checklist, and longitudinal studies using the JIBI Cohort checklist ^19^. Results of these checklists are presented in Appendix 2 in the standard risk of bias format.

### Data presentation and analyses

We summarized outcomes narratively. For figures without numbers, we extracted numbers using an online software (https://apps.automeris.io/wpd/). We describe interventions and outcomes based on the information provided in the studies. Median prevalence rates were presented as box-and-whisker plots. We decided not to perform a quantitative summary of the associations between the various correlates and mental health factors, due to a combination of heterogeneity in assessment measures and lack of control groups, and an overarching lack of descriptions necessary to confirm sufficient homogeneity. Our included studies not only varied greatly from one another, they most often did not report sufficient information regarding inclusion criteria, population, setting, and exposure to assess potential clinical heterogeneity. We graded the certainty of the evidence using the GRADE approach (Grading of Recommendations Assessment, Development, and Evaluation) ^20^.

## Results

### Results of the literature search

As of 11 May 2020, the *Live map of covid-19 evidence* project had screened 20,738 studies for covid-19 relevance, and categorized all studies with empirical data. We identified 557 studies coded to the topic *Experiences*, and 314 coded to *Healthcare workers*. Our database keyword search identified a further 218 relevant studies. Of a total of 1089 identified studies, 59 met our inclusion criteria for this systematic review ^8, 21-78^.

### Description of studies

Fifty-nine studies were included. Table 1 displays their summarized characteristics, while Appendix 3 displays characteristics of the individual studies. Thirty-nine studies were conducted in or included participants from China; four in Iran; three in the USA; two each in France, India, and Singapore; and one each from Australia, Germany, Italy, Malaysia, and Taiwan. Two studies reported results of international online surveys; one included respondents from 30 countries and the other from 91 countries. The majority of studies (47) were cross-sectional surveys, four were other cross-sectional designs; two studies reported surveys administered twice over time; three were qualitative studies; and one study searched within a database of existing online surveys. We also identified two systematic reviews ^29, 31^, which identified five primary studies ^8, 35, 39, 47, 76^.

**Table 1:** Summary of study characteristics.

| **Table 1: Summary of study characteristics** |  |  |
| --- | --- | --- |
|  | **N**<br>**(59 total)** | **%** |
| <u>Country (multiple allowed)</u> |  |  |
| Australia | 1 | 1.7% |
| China | 39 | 66.1% |
| France | 2 | 3.4% |
| Germany | 1 | 1.7% |
| Italy | 1 | 1.7% |
| India | 2 | 3.4% |
| Iran | 4 | 6.8% |
| Malaysia | 1 | 1.7% |
| New Zealand | 1 | 1.7% |
| Singapore | 2 | 3.4% |
| Taiwan | 1 | 1.7% |
| USA | 3 | 5.1% |
| Not applicable (systematic reviews) | 2 | 3.4% |
| Not applicable (international survey) | 2 | 3.4% |
| <u>Study design</u> |  |  |
| Survey | 47 | 79.7% |
| Other cross-sectional | 4 | 6.8% |
| Cohort/longitudinal | 2 | 3.4% |
| Qualitative | 3 | 5.1% |
| Systematic review | 2 | 3.4% |
| Other | 4 | 6.8% |
| <u>Healthcare setting (multiple allowed)</u> |  |  |
| Hospital | 42 | 71.2% |
| Specialist health services | 2 | 3.4% |
| Other | 3 | 5.1% |
| Not specified | 21 | 35.6% |
| <u>Population (multiple allowed)</u> |  |  |
| Allied health care workers | 3 | 5.1% |
| Clinical administration | 8 | 13.6% |
| Doctors | 33 | 55.9% |
| Emergency staff | 1 | 1.7% |
| Medical students | 2 | 3.4% |
| Nurses | 31 | 52.5% |
| Other | 13 | 22.0% |
| Not specified | 19 | 32.2% |
| <u>Exposure/intervention (multiple allowed)</u> |  |  |
| Frontline | 40 | 67.8% |
| Not frontline | 26 | 44.1% |
| Not specified | 10 | 16.9% |
| Intervention aimed at mental health | 6 | 10.2% |
| Other | 2 | 3.4% |

The studies reported on healthcare workers working in different settings: 43 studies reported on health care workers in hospitals, two studies were conducted in specialist health services outside hospitals, and three studies in other settings, while 21 studies did not specify the healthcare setting or only partially described multiple settings. No studies reported on nursing homes or primary care settings. In 40 studies, participants were frontline workers, while 26 studies reported on non-frontline workers. Frontline or non-frontline activities were unclear in ten studies.

Six studies reported on interventions to reduce mental health problems.

More than half of the studies included nurses (31) and/or doctors (33). Studies reported on a total of 54,707 healthcare workers, ranging from a case study with three participants to a survey of 11,118 participants.

### Methodological quality assessment of included studies

Appendix 2 displays the methodological quality assessments of individual studies. Overall assessments are displayed in Appendix 3, the description of included studies. Twenty-five studies were assessed as having low methodological quality (including eleven of 17 cross-sectional studies that provided only prevalence data), twelve medium, and sixteen high. The most common methodological weaknesses across all studies arose from insufficient reporting: samples, settings, and recruitment procedures were often not described thoroughly. While both systematic reviews had low scores on the AMSTAR, all three qualitative studies were assessed on the CASP checklist as valuable. Four studies had designs that we did not assess for quality: Jiang et al. ^56^, and Schulte et al. ^21^ reported on the development or uptake of mental health interventions; Liu et al. surveyed mental health questionnaires available online in China as of 8 February 2020 ^52^; and Martin presented three short case histories ^26^.

### Mental health interventions

Six studies reported on the implementation of interventions to prevent or reduce mental health problems caused by the covid-19 pandemic among healthcare workers. These interventions can be loosely divided into those targeting organizational structures, those facilitating team/collegial support, and those addressing individual complaints or strategies.

Two interventions involved organizational adjustments. The first intervention was reported on by two studies ^70, 72^. Hong et al.^70^ called it a “comprehensive psychological intervention” for frontline workers undergoing a mandatory two-week quarantine in a vocational resort, following two- to three-week hospital shifts. The quarantine itself was also described as part of the intervention, explicitly intended “to alleviate worries about the health of one’s family”. Other elements included shortened shifts; involvement of the labor union to provide support to healthcare workers’ families; and a telephone-based hotline that allowed healthcare workers to speak to trained psychiatrists or psychologists. This hotline had already been available to healthcare workers for four hours per week prior to the pandemic, but was made available for twelve hours, seven days a week. Chen et al.^47^ reported a second intervention that attempted to address individual complaints and facilitate collegial support. A telephone hotline was set up to provide immediate psychological support, along with a medical team that provided online courses to help healthcare workers handle psychological problems, and group-based activities to release stress. However, uptake was low, and when researchers conducted interviews with the healthcare workers to understand this, healthcare workers reported needing personal protective equipment and rest, not time with a psychologist. They also requested help addressing their patients’ psychological distress. In response, the hospital developed more guidance on personal protective equipment, provided a rest space, and provided training on how to address patients’ distress.

Schulte et al.^21^ targeted collegial support and building individual strategies through one-hour video “support calls” for healthcare workers called in from their homes, to describe the impact of the pandemic on their lives, to reflect on their strengths, and to brainstorm coping strategies. This intervention was implemented as a response to the hospital redeploying pediatric staff to work as covid-19 frontline staff, and reorganizing pediatric space to accommodate more pediatric and adult covid-19 patients.

The remainder of the interventions focused on individual complaints or strategies. Chung et al.’s^69^ intervention was an online questionnaire available through a hospital mobile phone application that allowed healthcare workers to request psychological support from a psychiatric nurse, and to fill out a short depression screening measure. Jiang et al.’s^56^ intervention began as an onsite, in-person psychological crisis intervention, in which psychiatrists and psychologists provided psychological care to healthcare workers. After in-person care was recognized as a transmission risk to the psychiatrists and psychologists, the intervention was developed to allow for remote provision.

### Changes in mental health during the pandemic

None of the studies that implemented mental health interventions reported on the effects of the interventions on healthcare workers. The only data available to approximate the impact of the pandemic on the mental health of Healthcare workers come from two longitudinal survey studies reporting on changes over time, both of low methodological quality.

Lv et al.^22^ surveyed healthcare workers before and during the outbreak, reporting no further information about the timeline. The study included both those working on the frontline and those with unclear exposure to covid-19. However, it is unclear whether respondents were the same at both time points. The prevalence of anxiety, depression, and insomnia increased over time, whether mild, moderate, moderate to severe, or severe (see Figure 2). During the outbreak, one out of every four healthcare workers reported at least mild anxiety, depression, or insomnia.

**Figure 1:**
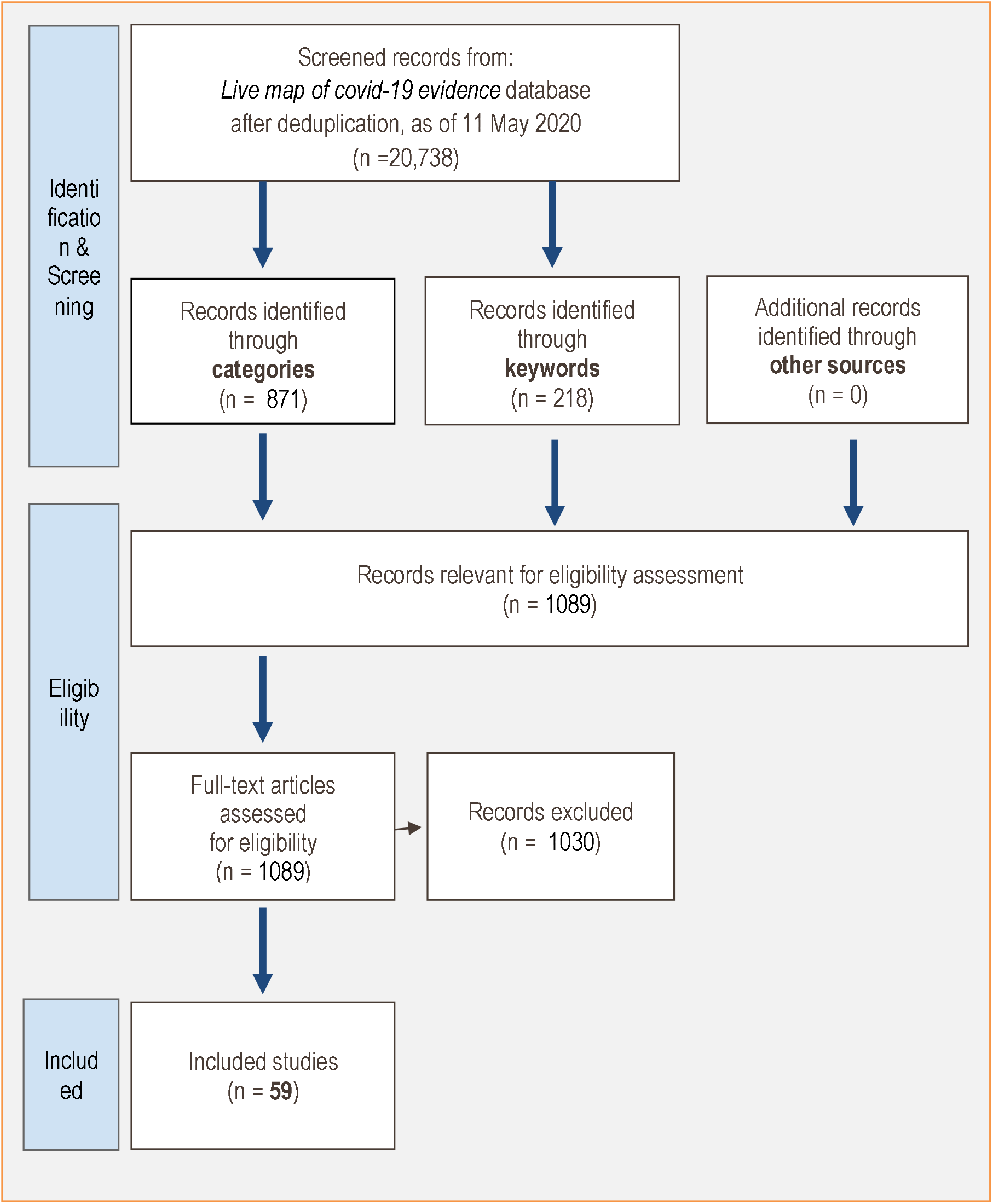
Live evidence map flow diagram of study inclusion.

**Figure 2:**
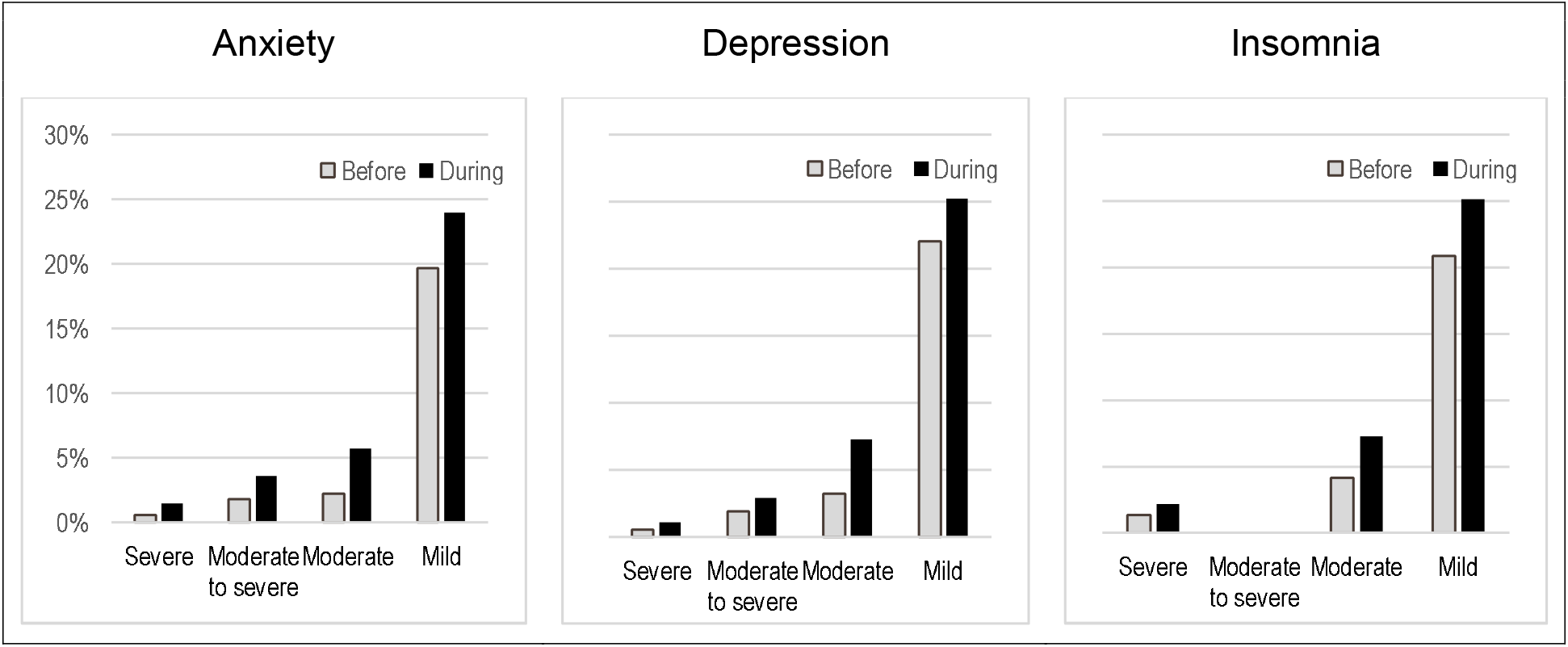
Anxiety, depression, and insomnia before and during the pandemic among Lv et al.’s sample of 8,028 Chinese healthcare workers before and during the pandemic.

Yuan et al.^27^ also administered a survey twice to 939 respondents during the pandemic (in February 2020), with two weeks between the surveys, and no attrition reported. Each respondent answered the same questions: “*I feel worried, I feel anxious, fidgeting and not knowing what to do, I feel frightened, I feel nervous and uneasy, I don’t think I can succeed even if I try hard*, and *I’ve been smoking or drinking a lot lately*.” The authors presented the changes per item after two weeks, rather than answers at both time points, and the answer scale was not reported. Worry worsened for 30% of participants, anxiety for 12%, fidgeting for 9%, fear for 15%, feeling nervous and uneasy for 13%, not thinking one can succeed for 4%, and an increase in smoking and drinking for only 1%. The proportion reporting improvement was similar for fidgeting, fear, and feeling nervous and uneasy, and more improved in not thinking one can succeed and for a reduction in smoking and drinking.

Two cross-sectional studies reported healthcare workers’ self-reported changes in mental health; both were also of low methodological quality due to insufficient reporting. In Benham et al.^78^, twelve Iranian psychiatry residents were re-deployed to work one frontline shift. Half of the residents reported that they experienced more distress after this shift. Abdessater et al.^28, 32^ studied 275 urology residents not working on the frontline. When asked to report the level of stress caused by covid-19, 56% reported a medium to high amount of stress, and the remaining reported none to low. Less than 1% had initiated a psychiatric treatment during the pandemic.

A third cross-sectional study^64^, also of low methodological quality, surveyed 60 healthcare workers in China in February, during the “outbreak period”. A different cohort of 60 healthcare workers were surveyed in March, during the “non-epidemic outbreak period”. The healthcare workers in to the second phase of the survey reported less symptoms of anxiety and depression, and higher health-related quality of life.

### Prevalence of mental health problems, and risk and resilience factors

Twenty-nine studies reported prevalence data of mental health variables as proportions or percentages. (Seventeen additional studies reported data as average scores on various instruments, and we did not extract this data.) We present box-and-whisker plots in Figure 3 to show the distribution of anxiety, depression, distress, and sleeping problems among the healthcare workers investigated in the 29 studies, using the authors’ own methods of assessing these outcomes

**Figure 3:**
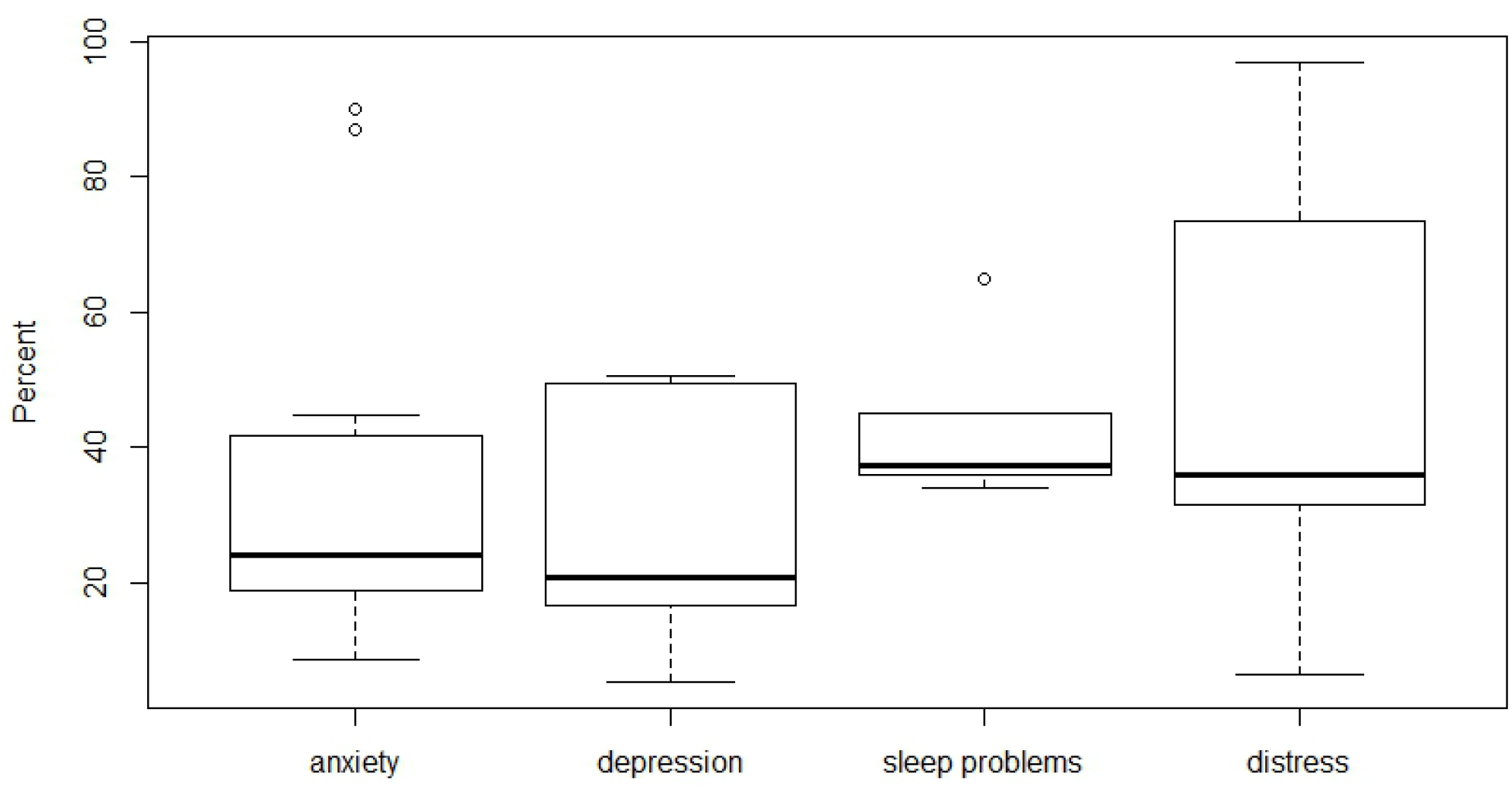
Boxplots of prevalence of anxiety, depression, sleep problems, and distress.

For anxiety, there were data from 22 studies. The percentage of healthcare workers with anxiety ranged from 9-90% with a median of 24%. For depression, there were data from 19 studies. The percentage with depression ranged from 5-51%, with a median of 21%. For sleep problems, there were data from six studies. The percentage with sleeping problems ranged from 34-65%, with a median of 37%. For distress, there were data from 13 studies. The percentage with distress ranged from 7-97%, with a median of 37%. Only one study^65^ reported prevalence of somatic symptoms, including decreased appetite or indigestion (59%) and fatigue (55%).

The summary of findings table below displays median prevalence rates across the studies contributing to each mental health outcome.

Our confidence in the reported results of levels of anxiety, depression, distress and sleep problems in health care workers during the covid-19 pandemic was assessed using the GRADE approach to be very low. Our confidence was reduced (downgraded), as shown in Table 2, due to high risk of bias, large heterogeneity and imprecision.

**Table 2:**
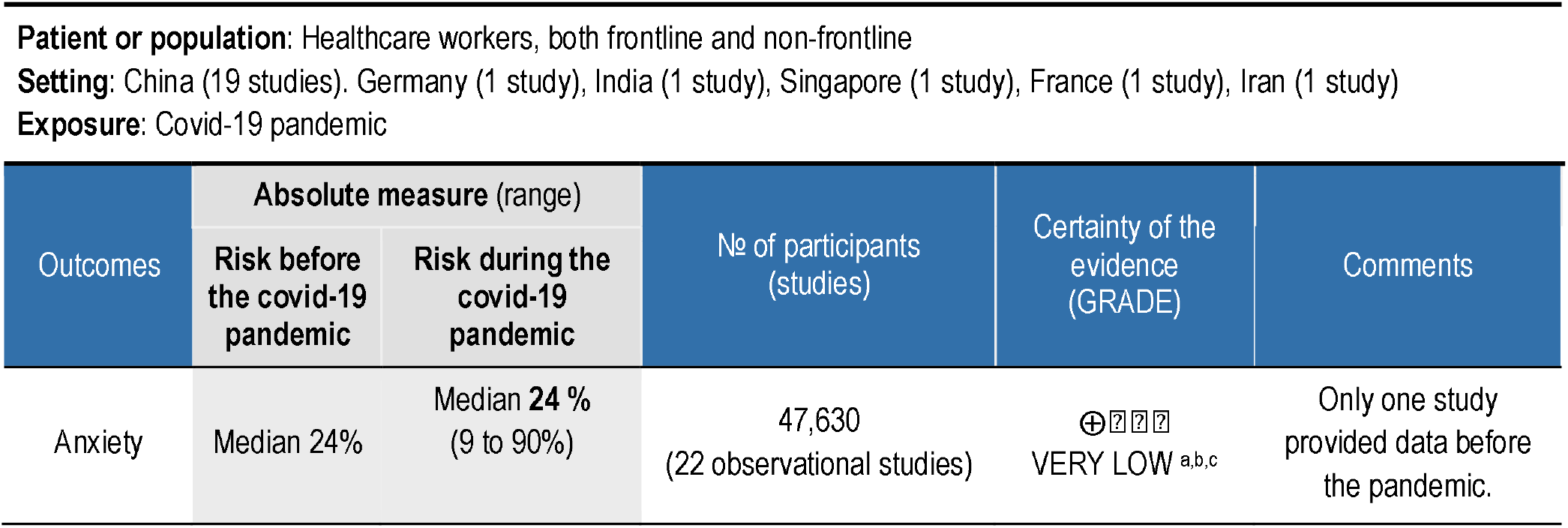
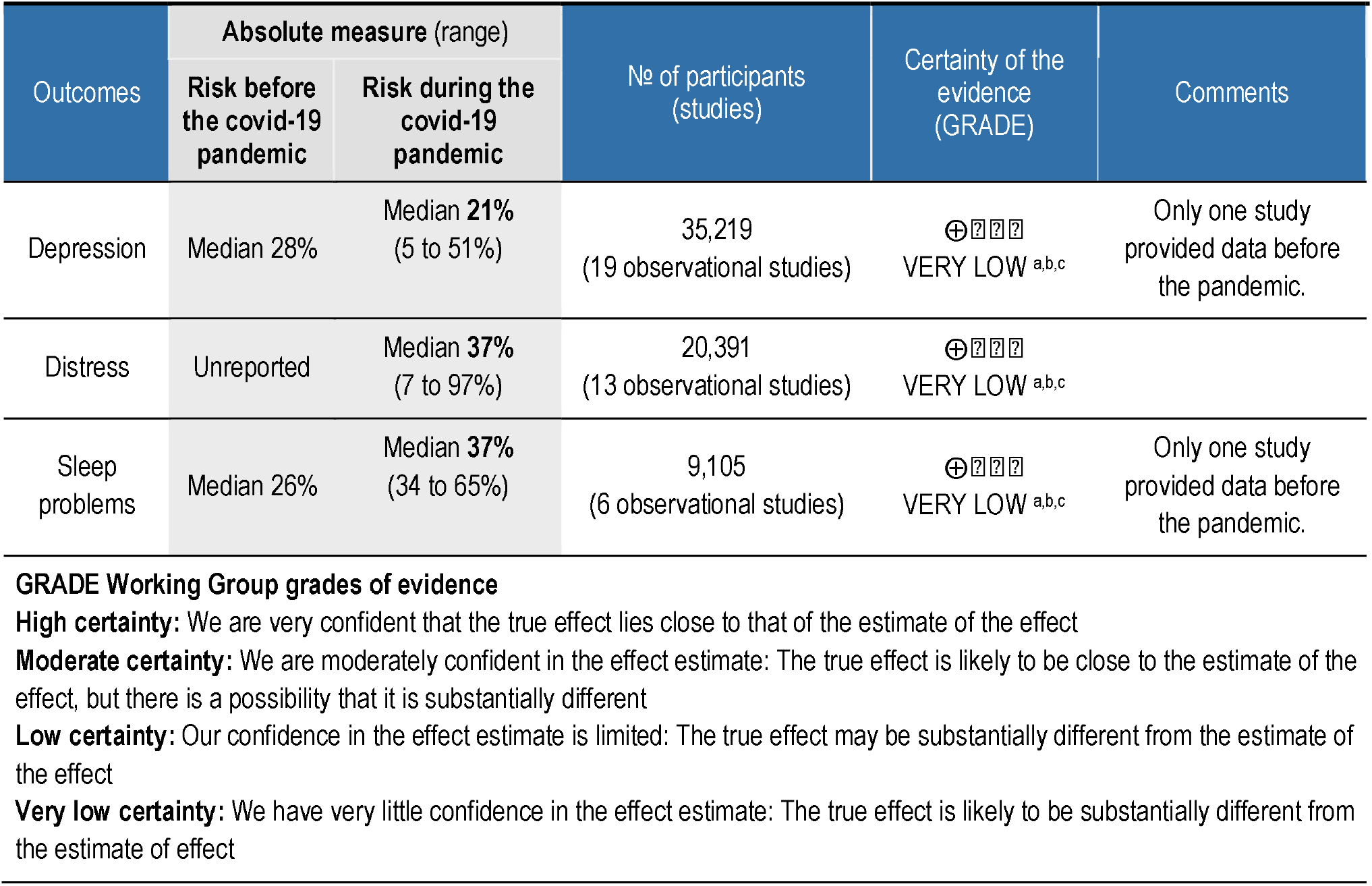
Summary of findings table.

Twenty-two studies reported one or more variables associated with mental health problems in health care workers during the pandemic. The most common risk factors correlated with increased risk of mental health problems were exposure to covid-19 patients ^28, 39, 46, 49, 60, 63, 74^, being female^8, 25, 39, 46, 54, 60^, and worry about being infected ^25, 46, 58, 63^. In three studies, worrying about family members being infected was a risk factor^42, 58 62^. The 22 studies mentioned a number of other factors once each, that we do not report here.

The most commonly reported protective factor associated with reduced risk of mental health problems was having social support^35, 49, 53, 74^. Two studies directly measured self-perceived resilience. Bohlken et al.^38^ asked their sample of psychiatrists and neurologists to assess how resilient they were on a Likert scale from 1-5 (“not applicable” to “completely applicable”), and 86% selected the two highest categories. Cai et al. ^34^ compared experienced frontline workers with inexperienced frontline workers, and found that inexperienced workers scored lower on total resilience on the Connor-David resilience scale as well as within each of three subscales, and had more mental health symptoms. Inexperienced workers were also younger and had less social support available to them.

### Strategies and resources used

Ten studies reported that healthcare workers utilized other resources or had individual strategies to address their own mental health during the pandemic, separate from formal interventions.

Six studies reported healthcare workers’ utilized support from family/friends during the pandemic. “Family” was the most common stress coping mechanism utilized by Louie et al.’s^42^ sample (78.5%). Sixty-five percent of Sun et al.’s^67^ sample sought social support to relieve stress. Forty-three percent of Cai et al.’s^58^ sample rated social support from friends and family as a “very important” strategy, on a scale from “not at all” to “very important”; a similar proportion of Louie et al.’s sample said they used telecommunication with friends as a coping mechanism (43.8%). Cao et al.^72^, reporting themes from interviews, wrote that telecommunication with family members was the most frequently utilized coping mechanism, while a majority identified talking with friends as important. Mohindra et al.’s^45^ narrative report of an unreported amount of interviews also identified support from family and colleagues as a main emotional motivational factor for healthcare workers to continue working.

Professional and informal help were strategies reported by two studies each. A minority of healthcare workers in Cai et al.^58^ said that seeking help from a psychologist was important. Counselling, therapy, or other professional interventions were used by 18-36% of Kang et al.’s^43^ and Liu et al.’s^48^ samples, respectively. Half of Kang et al.’s sample used psychological resources available through the media, and 36% used other psychological materials. Less than one third of Zhang et al.’s^74^ sample reported that psychological support from news or social media was helpful, although the amount who utilized news or social media was not reported.

In Sun et al.’s^67^ qualitative study of nurses’ psychological experience of treating patients with covid-19, coping and self-care styles was an emergent theme, and subsequently described quantitatively. All nurses utilized active psychological defense mechanisms (such as mindfulness) or more passive strategies (such as distraction). Seventy percent made “life adjustments” such as sleeping, exercising, or eating more. Sixty-five percent sought social support for stress relief. Just under half (45%) sought or used external information to adjust their thought patterns.

### Perceived need and preferences for interventions aimed at preventing or reducing negative impact on mental health

Utilization of formal and informal strategies can be interpreted as an indicator of healthcare workers’ preferences. Seven studies asked healthcare workers directly about whether they needed mental health help, and their preferences regarding such help.

Several studies reported a low level of interest in professional psychological services. Five percent in Cao et al.^72^ explicitly said they would not want professional help. Similarly, Guo et al.^57^ asked their sample “how to deal with psychological distress”, and only 14% selected psychological counselling. The majority said distress could be endured or solved individually, and a majority also said talking to friends or family could help. Nineteen percent said online information could help.

Kang et al.^43^ found slightly higher levels of interest in professional resources. When asked from whom they prefer to receive “psychological care” or “resources”, 40% answered psychologists or psychiatrists, 14% answered family or relatives, 15% answered friends or colleagues, 2% answered others, and 30% said they did not need help. The authors found that preferred sources of psychological resources were related to the level of psychological distress. In a structural equation model that uncovered clusters of healthcare workers with different distress levels (subthreshold, mild, moderate, and severe), those with moderate and severe distress more often preferred to receive care from psychologists or psychiatrists, while those with subthreshold and mild distress more often preferred to seek care from family or relatives.

In two studies, participants specified that they had a greater need for personal protective equipment than for psychological help. Chung et al.^69^ reported this in a survey that allowed healthcare workers to describe their needs and concerns in free text and to request contact with a psychiatric nurse. While 3% requested such contact, nearly half of those who answered the free text question about their psychiatric needs wrote that they needed personal protective equpiment instead, and 20% said they were worried about infection. Chen et al.’s ^47^ study was to understand why uptake of their psychological intervention was so low, and findings were identical to Chung et al.’s: “Many staff mentioned that they did not need a psychologist, but needed more rest without interruption and enough protective supplies” (p. e15).

Only one study explored how healthcare workers would be willing to provide mental health services to other healthcare workers: twelve psychiatry residents were re-deployed as frontline workers for one shift in Benham et al.’s^78^ study. After that shift, none were willing to provide face-to-face mental health services to other healthcare workers, although 75% said they would provide online services. They identified healthcare workers of deceased patients as possible target populations for online services.

### Experience and understanding of mental health and related interventions

Three qualitative studies assessed as valuable were included. Two interconnected themes across all three studies were distress stemming both from concern for infecting family members, and from being aware of family members’ concern for the healthcare workers.

Wu et al.^71^ explored reasons for stress during interviews with healthcare workers at a psychiatric hospital. While these healthcare workers were not on the frontline, they felt they were at higher risk of exposure than healthcare workers at a general hospital. Their wards were crowded, and several patients were admitted from emergency rooms with aggressive behaviors that made social distancing difficult or that posed direct challenges to healthcare workers’ use of personal protective equipment (such as tearing masks). Healthcare workers felt unprepared because psychiatric hospitals had no plans in place. At the same time, they also felt that their peers on the frontline were providing more valuable care. An additional source of stress was knowledge of their own risk of infection and transmission to family members, particular to elderly parents in their care, and to children who were at home and whose schoolwork had to additionally be managed. The disruption of the pandemic to nurses’ personal lives and career plans was another stressor.

Sun et al.^67^ interviewed twenty frontline nurses about their psychological experiences of frontline work. Similar themes as Wu et al.’s sources of stress were reported, particularly the fear of infecting friends and family. Elderly parents and children at home were again mentioned, and concern was great enough that several respondents did not tell their family they were working on the frontline, while others did not live at home during this period. As with Wu et al.’s non-frontline workers, these healthcare workers also reported fear and anxiety of a new infectious disease that they felt unprepared to handle on a hospital-level, unprepared to treat on a patient-level, and from which they were unable to protect themselves. The first week of training and the first week of actual frontline work was characterized by these negative emotions, which were then joined – not necessarily replaced – by more positive emotions such as pride at being a frontline nurse, confidence in the hospital’s capacity, and recognition by the hospital.

Yin et al.^73^ used a framework of existence, relatedness, and growth theory to analyze nurses’ psychological needs. They reported nurses’ identification of *existence* needs as primarily health and security: their own physical and mental health, personal protective equipment, and emotional stability for their family. Their need for *relatedness* was represented by needs for relationships and affection, as well as for care, help, and support from colleagues and bosses, as well as from outside the hospital. Finally, *growth* needs referred to needing knowledge of covid-19 infection prevention and control, particularly from the authorities.

Mohindra et al.’s^45^ cross-sectional survey also reported experiences of mental health promotion narratively, with similar results as Yin et al: more knowledge of covid-19 could strengthen motivation, as could emotional support. Affecting them negatively were fears of infecting their families, particularly because their families would suffer more financially from needing to be quarantined than they already were suffering under the lockdown; fears of using personal protective equipment incorrectly; and feeling unequipped to handle patients’ non-medical needs. Healthcare workers reported that stigma suppressed patients’ provision of accurate travel and quarantine history. This was an issue they were ill-equipped to help patients address when they returned to the community. Healthcare workers also reported that they were stigmatized, because they were potential sources of infection.

## Discussion

This systematic review identified 59 heterogeneous studies that examined the mental health of healthcare workers during the covid-19 pandemic. The total of 54,707 participants included mainly frontline nurses and physicians, but also other healthcare workers who provided clinical care, administration, or other clinical tasks. Studies reported a variety of outcomes and situations, including the implementation of interventions to prevent or reduce mental health problems, other resources and strategies utilized by healthcare workers, and on healthcare workers’ mental health responses to re-deployment as frontline workers. While the majority of studies were cross-sectional and assessed as having high risk of bias, several patterns in their findings were evident: more healthcare workers were interested in social support to alieve mental health impacts, only a minority were interested in professional help for these problems, and yet interventions described in the literature largely seemed to focus on relieving individual symptoms. The current study reveals a mismatch between the likely organizational sources of psychological distress, such as workload and lack of personal protective equipment, and how healthcare systems are attempting to relieve distress at an individual level.

Between one and two of every five healthcare worker reported anxiety, depression, distress, and/or sleep problems. Only one study reported on somatic symptoms such as changes in appetite. These findings comport with much of the existing literature; healthcare workers in general, and particularly intensive care nurses and physicians, are known for elevated levels of distress compared to the general population ^79-83^. Findings from the two studies following healthcare worker over two timepoints during the pandemic indicate that these complaints increased from the first timepoint to the next. Thus, there is reason to believe that the pandemic and working conditions during the pandemic negatively affects healthcare workers, although more longitudinal studies are needed to confirm this hypothesis.

There are many plausible mechanisms. While our included studies do not allow us to draw any conclusions regarding causality, their findings – particularly perceived need and preferences – can point us in certain directions. First, high workload and the absence of healthy rotation schedules that accommodate adequate rest, sleep, and restoration over time may have contributed to the mental health problems reported by studies. Sleep problems and insomnia in particular are likely mediators of psychological distress^84^. Both the qualitative and the cross-sectional studies identified exposure to patients with covid-19 and/or a lack of personal protective equipment and subsequent fear of infecting colleagues, family, friends, and oneself as major contributors to the distress reported by healthcare workers. Even when personal protective equipment was available, not all healthcare workers felt trained enough for proper use, an example of a discrepancy between the demands of a job and the skills possessed^85^, which itself is a well-known stressor for healthcare workers in non-pandemic times. The escalation of work-related pressure, rotation of healthcare workers to the frontline, new tasks, and related increases in assignments during crises or disasters such as this pandemic are a recipe for occupational stress, unless handled appropriately by hospitals.

Most formal interventions implemented to prevent or relieve mental health problems focused not on organizational factors or on collegial factors, but on individual symptoms. They tended to do so by facilitating the provision of individual mental health services to healthcare workers. The underlying focus of these interventions appeared to be individual psychopathology, without further systematic exploration of the impact of organizational or collegial factors on adverse mental health outcomes. The focus on individual risk and resilience factors and pathology in research may hinder the discovery of underlying organizational faults, which could be more appropriate targets of intervention. This focus on the individual rather than system-level factors is also common in interventions for healthcare worker burn-out before the pandemic^86^. The most striking illustration of this was the finding shared by two studies^47, 69^ that healthcare workers said personal protective equipment would benefit their mental health more than professional help. On the other hand, it is possible that Healthcare workers could benefit from professional mental health interventions more than they recognize or report, and that under-recognition is related to occupational culture, fear of stigma or weakness, or simply cultural differences, as the two studies in question both reported on Chinese healthcare workers.

The possible risk and protective/resilience factors reported by our included studies are similar to those identified in other recent reviews of healthcare workers’ mental health during other novel viral outbreaks such as SARS, MERS, Ebola, and H1N1. These factors, not related to individual psychopathology, could be areas for healthcare settings to proactively address: junior status, higher exposure, longer quarantine time, having an infected family member, lack of practical support, stigma, and younger age were risk factors of distress in Kisely et al’s^87^ review. De Brier et al.^88^ also reported exposure, quarantine, and health fear as risk factors. Protective factors identified in these two reviews were similar: clear communication, access to adequate personal protective equipment, adequate rest, and both practical and psychological support in Kiseley et al.; clear communication and support from the organization, social support, and a personal sense of control in De Brier et al.

Reported strategies and resources are an important finding of this review: seeking social contact and support was the most common strategy reported by healthcare workers to take care their own mental health, and there was less interest or utilization in professional mental health services. At the same time, there are likely barriers to availing themselves of existing social support during the pandemic. High work burdens combined with healthcare workers’ fear of infecting others and high levels of worry may prevent them from accessing or seeking existing social support. Healthcare workers’ own psychological reactions to these situations, such as distress or irritation, may lower the empathy and support extended to them from social networks. Accessing and capitalizing on such support could be another appropriate target of an intervention, as in Schulte et al^21^. A strength of this review is its depth; it is the most comprehensive review to date of the mental health of healthcare workers under the covid-19 pandemic. Our quality assessment of qualitative and quantitative studies should help other researchers in the evidence synthesis process, if they wish to use methodological quality in their inclusion criteria. We followed the Norwegian Institute of Public Health’s rigorous methodological standards for systematic reviews, such as two researchers screening and assessing eligibility. An additional methodological strength is our utilization of the *Live map of covid-19 evidence*, one of the first reviews to do so (see also two reports^89, 90^ and one diagnostic accuracy study^91^. By using our map, we quickly identified 871 studies that had already been categorized to our topic and population of interest, without having to search in academic databases and screen again.

While not being able to conduct a meta-analysis is unfortunate, it was appropriate not to assume that poorly reported studies were homogenous enough. The principle of homogeneity tends to be overlooked by systematic reviewers eager to produce a summary estimate, but if met, means that all studies included were similar enough that their participants can be considered participants of one large study^92^. The result, however, is that the prevalence data about mental health problems does not provide a summary estimate that can be generalized. Other weaknesses are those common to rapid reviews due to time pressure, such as fewer details about the included studies’ populations being presented than normally reported.

The covid-19 pandemic has resulted in a flood of studies, many of which have been pushed through the peer-review process and published at speeds hitherto unseen (see Glasziou^93^ for a discussion). It is therefore not surprising that the majority of our included 59 studies were assessed as having a high risk of bias or being of low methodological quality. Lack of information on samples or procedures was a common limitation, leading to serious implications to the generalizability and validity of findings. We also call on journals and researchers to balance the need for rapid publication with properly conducted studies, reviews and guidelines^94^.

## Conclusion

Healthcare workers in a variety of fields, positions, and exposure risks are reporting anxiety, depression, distress, and sleep problems during the covid-19 pandemic. Causes vary, but for those on the frontline in particular, a lack of opportunity to adequately rest and sleep is likely related to extremely high burdens of work, and a lack of personal protective equipment or training may exacerbate mental health impacts. Provision of appropriate personal protective equipment and work rotation schedules to enable adequate rest in the face of long-lasting disasters such as the covid-19 pandemic seem paramount. Over time, many more healthcare workers may struggle with mental health and somatic complaints. The six studies exploring mental health interventions mainly focused on individual approaches, most often requiring healthcare workers to initiate contact. Proactive organizational approaches could be less stigmatizing and more effective, and generating evidence on the efficacy of interventions/strategies of either nature is needed. As the design of most studies was poor, reflecting the urgency of the pandemic, there is also a need to incorporate high-quality research in disaster preparedness planning.

## Data Availability

Secondary data analyzed in this review may be available upon reasonable request.

## Author Contributions

GEV conceived of this review and conducted the GRADE assessments. SS and SØS wrote the first drafts of the introduction and discussion. AEM identified the studies within the map for this review. JPWH and EVH wrote the first draft of the methods. AEM, SV, GS, GEV, SF, EVH, and JPWH developed the methods of the *Live map of covid-19 evidence*. SØS contributed to identifying outcomes. AEM, SF, and GEV extracted data and assessed study quality. AEM and GS conducted the analyses. All authors contributed to the protocol and design of this review. All authors have read and approved the final draft of this manuscript.

## Declarations

The corresponding author has the right to grant on behalf of all authors and does grant on behalf of all authors, an exclusive license (or non exclusive for government employees) on a worldwide basis to the BMJ Publishing Group Ltd to permit this article (if accepted) to be published in BMJ editions and any other BMJPGL products and sublicenses such use and exploit all subsidiary rights, as set out in our license.

### Competing interest

All authors have completed the Unified Competing Interest form (available on request from the corresponding author) and declare: no support from any organization for the submitted work; no financial relationships with any organizations that might have an interest in the submitted work in the previous three years; no other relationships or activities that could appear to have influenced the submitted work.

Transparency: The lead author affirms that the manuscript is an honest, accurate, and transparent account of the study being reported; that no important aspects of the study have been omitted; and that any discrepancies from the study as planned (and, if relevant, registered) have been explained.

### Ethical approval

No ethical approval was required for this systematic review.

### Patient and public involvement

This rapid systematic review did not seek involvement of the population of interest (healthcare providers) due to time constraints. Healthcare providers will instead receive direct dissemination of results.

